# A large decrease in the magnitude of seasonal fluctuations in mortality explains part of the increase in longevity in Sweden during 20th century

**DOI:** 10.1101/2020.04.10.20060780

**Authors:** Anders Ledberg

## Abstract

**BACKGROUND:** Mortality rates are known to depend on the seasons and, in temperate climates, rates are highest during winter. The magnitude of these seasonal fluctuations in mortality has decreased substantially in many countries during the 20th century, but the extent to which this decrease has contributed to the concurrent increase in life expectancy is not known. Here, I describe how the seasonality of all-cause mortality among people ages 60 years or more has changed in Sweden between 1860 and 1995, and investigate how this change has contributed to the increase in life expectancy observed during the same time period.

**METHODS:** Yearly sex-specific birth cohorts consisting of all people born in Sweden between 1800 and 1901 who reached at least 59 years of age were obtained from a genealogical database. The mortality rates for each cohort were modeled by an exponential function of age modulated by a sinusoidal function of time of year. The potential impact of seasonal fluctuations on life expectancy was investigated by a novel decomposition of the total mortality rate into a seasonal part and a part independent of the seasons. Cohort life expectancy at age 60 was used to quantify changes in lifespan during the time period.

**RESULTS:** The magnitude of seasonal fluctuations in mortality rates decreased substantially between 1860 and 1995. For cohorts born in 1800, the risk of dying during the winter season was almost twice that of dying during summer. For cohorts born in 1900, the relative increase in winter mortality was 10%. Cohort life expectancy at age 60 increased by 4.3 years for men and 6.8 years for women, and the decrease in seasonal mortality fluctuations accounted for approximately 40% of this increase in average lifespan.

**CONTRIBUTION:** By following a large number of extinct cohorts, it was possible to show how the decrease in seasonal fluctuations in mortality has contributed to an increase in life expectancy. The decomposition of total mortality introduced here might be useful to better understand the processes and mechanisms underlying the marked improvements in life expectancy seen over the last 150 years.

## 1 Introduction

Mortality rates in humans depend on the seasons – in countries with temperate climate the rate of dying is typically higher during winter months (e.g., Tabellkommissionen, 1857; Rosenwaike, 1966; Healy, 2003; Davie et al., 2007; Rau, 2007; Fowler et al., 2015). Such seasonal fluctuations in mortality rates, with a peak during winter, is seen for most major causes of death but the magnitude varies between different causes. For example, mortality from cardiovascular diseases show larger seasonal fluctuations than mortality from cancers (e.g., Rosenwaike, 1966; Näyhä, 1980; Reichert et al., 2004; Parks et al., 2018; Rau, 2018). Other causes of death, such as accidents and suicides, might have very different dependencies on the seasons and have been reported to have higher rates during the summer (e.g., Näyhä, 1980; Parks et al., 2018).

In cross-sectional data, the seasonal variation in mortality differs between age groups and the winter peak in mortality is most pronounced among older people (e.g., Näyhä, 1980; van Rossum et al., 2001; Feinstein, 2002; Rau, 2007; Parks et al., 2018; Rau, 2018). For example, Parks and coworkers analyzed all deaths occurring in the United States between 1980 and 2016 and found that all-cause mortality was strongly dependent on the seasons in all age groups above 55 years; in younger age groups, on the other hand, the dependence on the seasons was weak or non-existent (Parks et al., 2018). This age-dependency in the seasonal patterning of mortality is likely a consequence of causes of death being differentially distributed by age. For example, accidents and injuries (external causes) constitute a much larger fraction of deaths among younger people. Indeed, 36% of all deaths in Sweden last year among people under age 55 were due to external causes, for deaths above 55 the corresponding fraction was 3.9%.

The seasonal variability of mortality rates is likely caused by multiple, and interacting, factors, the most prominent are infectious diseases of the respiratory system and their consequences (e.g., Anderson and Le Riche, 1970; Reichert et al., 2004), and direct and indirect effects of cold temperatures (e.g., Rogot and Padgett, 1976; Kunst et al., 1993; Curriero et al., 2002; Anderson and Bell, 2009). The former cause might be harder to protect against (c.f. Simonsen et al., 2007; Osterholm et al., 2012), but major improvements have been made during the last one hundred and fifty years in the ability to safely protect against the impacts of weather.

The magnitude and phase of seasonal mortality variations have not been constant over time (Rau, 2007). Indeed, in many countries there has been a marked decrease in magnitude during the 20th century (Näyhä, 1980; Keatinge et al., 1989; Kunst et al., 1991; Seretakis et al., 1997; Lerchl, 1998; Rau, 2007). During the same time period, life expectancy has increased substantially in most countries, but the extent to which the decrease in magnitude of seasonal fluctuations has contributed to this increase is not known. The increase in life expectancy at birth is a consequence of decreased mortality rates at all ages, and even if infant and child mortality rates have decreased most, in relative terms, the decrease in mortality rates at older ages has also contributed (e.g., Janssen et al., 2005; Statistics Sweden, 2020, Ch. 4).

The main aim of this paper is to investigate changes in seasonal fluctuations in all-cause mortality between 1860 and 1995 among the elderly in Sweden, and furthermore to estimate the impact these changes might have had on life expectancy. The time period analyzed in this work was chosen to cover a period during which mortality rates, and hence life expectancy, changed dramatically in Sweden. The focus on changes in mortality among elderly (aged 60 years or more) is motivated partly by that this group was likely more affected by extraneous conditions such as the weather, and by previous research showing that seasonality in this age group is present for many common causes of death (e.g., Feinstein, 2002; Parks et al., 2018).

Seasonality of all-cause mortality is studied by following sex-specific annual birth cohorts consisting of all people born in Sweden from 1800 to 1901 who survived at least until age 59 (c:a 4.7 million people in total). Cohort mortality rates are modeled by an exponentially increasing function of time with a super-imposed one-year periodicity. The model allows the seasonality of each cohort to be described by just two parameters, amplitude and phase, and changes in seasonality are quantified by comparing the estimated amplitudes. The contribution of seasonality to life expectancy is estimated by decomposing the total mortality rate into two parts: A *lower-bound mortality* that increases exponentially with age but does not vary with the seasons. This can be thought of as the mortality rates that would prevail under the optimal conditions of late summers. The other part of the decomposition, the *seasonal mortality*, depends on the seasons but not on age.

## 2 Methods

### 2.1 Data

Mortality data for people who were born and died in Sweden were obtained from the “Swedish Book of Deaths” issued by the The Federation of Swedish Genealogical Societies (Sveriges Släktforskarförbund, 2017). This is a database compiled from a range of official sources and contains information on times and places of births and deaths for people that have died in Sweden since 1860. The coverage is almost complete. Since there is no information about causes of deaths in this database this study is based on all-cause mortality.

Birth cohorts were formed for all years between 1800 and 1901, for men and women separately. The focus in this work is on all-cause mortality for those who survived until the beginning of the year in which they would have turned 60 years old, i.e., the they all survived at least until their 59th birthday. Since the database does not contain information about people who died before 1860, this age limit was needed in order to include birth cohorts from the entire 19th century. However, using a slightly different age limit, and thus a different range of birth years, only led to minor changes in the results (not shown). In total, there were almost 4.7 million people who fulfilled the inclusion criteria. Supplementary table S1 lists the number of people in each cohort.

### 2.2 Follow-up time

To investigate how mortality rates vary with the seasons, follow-up should be expressed in terms of calendar time. However, mortality rates also depend (strongly) on age, and it is therefore necessary to first express age (for each individual) in terms of calendar time.

Start of follow-up (i.e., time zero) was taken as the 1st of January the year cohort members would have turned 60 years old, and follow-up time was measured in days. Since all members are (by construction of the cohorts) alive at their 59th birthday, we can let the date of the 59th birthday be age zero. Let *b*_*i*_ denote the calendar day when cohort member *i* was born, expressed as the number of days since the beginning of the year. For example, for someone born the 13th of February, *b*_*i*_ = 30 + 13 = 43. The age of cohort member *i*, in days from time zero, *a*_*i*_(*t*), can then be expressed as a function of calendar time: *a*_*i*_(*t*) = 365 − *b*_*i*_ + *t*.

### 2.3 Model of mortality rates

Mortality rates for persons aged 60 years and above are often modeled by an exponential function of age (e.g., Gompertz, 1825; Olshansky and Carnes, 1997), and an exponential model has previously been shown to provide a good fit to a subset of the data used here (Ledberg, 2020). To investigate how mortality rates depend on the seasons, this exponential model was extended by including a (1-year) periodic component. Thus, with follow-up time *t* expressed in days, the mortality rates of persons born on day *b*_*i*_ and belonging to cohort *j, λ*_*j*_(*t*) say, are assumed to obey:

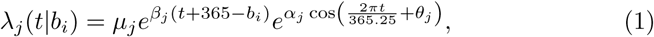

where *µ*_*j*_ > 0, *β*_*j*_ > 0, *α*_*j*_ ≥ 0, and *θ*_*j*_ are parameters that depend on the cohort (i.e., on sex and birth year). Note that, according to this model, the effects of the seasonal modulations, on a ratio scale, are assumed to be independent of age. This is a useful first approximation suitable for the current study (see Results), however, more complex models can easily be formulated within the framework outlined herein.

#### 2.3.1 Parameter estimation

To fit the model (Eq. 1) to data, an approximation related to the piecewise exponential model (Laird and Olivier, 1981) was used. Assume that time is measured in units of days, and consequently that *t* denotes the *t*-th day after beginning of follow-up. Note that this discretization of time implies that the mortality rate, *λ*(*t*), becomes piecewise constant. Let *N*_*t*_ denote the number of persons alive at the beginning of day *t* and *d*_*t*_ the number of persons who died on this day.^1^ As long as *λ*(*t*) << 1, *d*_*t*_ is well approximated by Poisson random variable with expected value *E*(*d*_*t*_) = *λ*(*t*)*N*_*t*_. Taking logarithms on both sides, using Eq. 1, and a well-known trigonometric identity for the cosine of the sum of two angles gives

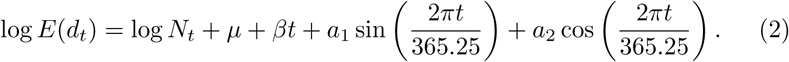

Since *d*_*t*_ is approximately distributed as a Poisson random variable, the parameters of Eq. 2 (and hence of Eq. 1) can be estimated by Poisson regression using existing software routines, for example the glm function in R. The amplitude *α* and phase *θ* in Eq. 1 are obtained from the following relations 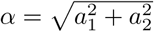 and *θ* = arctan(−*a*_1_*/a*_2_). In the supporting material it is shown that the parameter estimates obtained using Eq. 2 differ only marginally from those obtained by a direct maximization of the likelihood corresponding to Eq. 1. Estimation using Eq. 2 is preferred because it is faster and can be done using existing software. Data and code to reproduce the figures is available upon request.

The model was fit to data for each birth cohort separately, starting from the year where cohort members would have turned 60, and ending by the year they would have turned 95. This upper-age restriction was imposed because estimates of daily mortality rates are highly variable when *N*_*t*_ becomes small, and consequently hard to fit with any model. This restriction led to that less than two percent of all deaths in the database were censored.

### 2.4 Magnitude of seasonality

The magnitude of seasonality in a given cohort was quantified by fitting the model to data from the cohort and using the predicted ratio of the highest to lowest mortality rate in a given year. This can be interpreted as a risk ratio, approximating how many times higher the risk is of dying in the winter compared to dying in the summer. Note that according to the model (Eq. 1) this magnitude is constant for a given birth cohort, and consequently, this single number can be used to track how the magnitude of seasonality changed between cohorts. The ratio in cohort *j, h*_*j*_ say, is obtained from Eq. 1 as

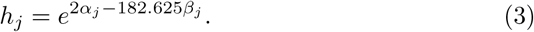

Pointwise approximate confidence intervals for *h*_*j*_ were obtained by first sampling values of *a*_1*j*_, *a*_2*j*_ and *β*_*j*_ from a multivariate normal distribution with mean and covariance matrix given by the fit of Eq. 2. These sampled parameters were then transformed according to Eq. 3, and confidence intervals were estimated from the empirical distribution so obtained. The normality assumption in this procedure is warranted by the fact that maximum likelihood estimates are asymptotically normal, and that the degrees of freedom in fitting the models were moderately large.

### 2.5 Partitioning the mortality

In order to investigate the effect of the seasonal changes in mortality on cohort life expectancy, Eq. 1 is recast in the following form (suppressing the dependency on cohorts of clarity)

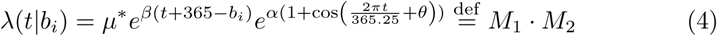

Where 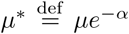. According to this formulation, the mortality rate at time *t* is partitioned into a first part, 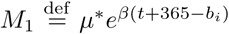, which is growing exponentially with age and represents the lowest achievable mortality for a given cohort. This part will be referred to as *lower-bound mortality*. The second part, 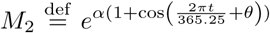, depends on the seasons but not on age, and will be referred to as the seasonal part of total mortality. Since the cosine function is bounded by *±*1 it follows that *M*_2_ ≥ 1, and thus acts to increase the mortality rate multiplicatively. Note that a change in mortality (between cohorts) can come about through a change in *M*_1_, *M*_2_, or in both.

### 2.6 Contribution to mean length of life

Cohort mortality rates *λ*(*t*) and life expectancy are related through well-known expressions, see for example Kalbfleisch & Prentice (Kalbfleisch and Prentice, 2002). The probability density of life times, *f* (*t*) say, is related to the mortality rate by

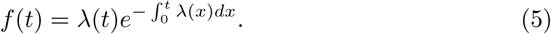

Note that time zero, in this work, was defined to be the first of January the year when cohort members would have turned 60, so the above expression actually represent a conditional density of life times. Mean life duration, conditional upon reaching first of January of the 60th life-year, is then obtained as the mean value of the corresponding distribution:

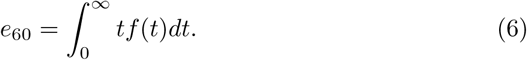

The integrals in Eq. 5 and Eq. 6 do not, in general, have closed form solutions and were therefore evaluated numerically.

Using the decomposition of Eq. 4 it is now possible to answer the following counterfactual question: “what would cohort life expectancy have been if mortality rates would have been given by the lower-bound mortality?” This is accomplished by evaluating Eq. 6 for *λ* = *M*_1_. The contribution of seasonality can then be obtained by evaluating Eq. 6 with *λ* = *M*_1_ *M*_2_ and taking the difference between these estimates.

#### 2.6.1 True average length of life

Since the cohorts are all extinct, life expectancy (conditioned on reaching any particular age) is simply given by the average over the cohort members who reached this age.

### 2.7 Visualization

To visualize seasonality in Figures 1 and 2, the time series of daily mortality rates were smoothed by a running-line smoother of 60 days width (i.e., each point is replaced by the local mean of the 60 surrounding points).

**Figure 1:**
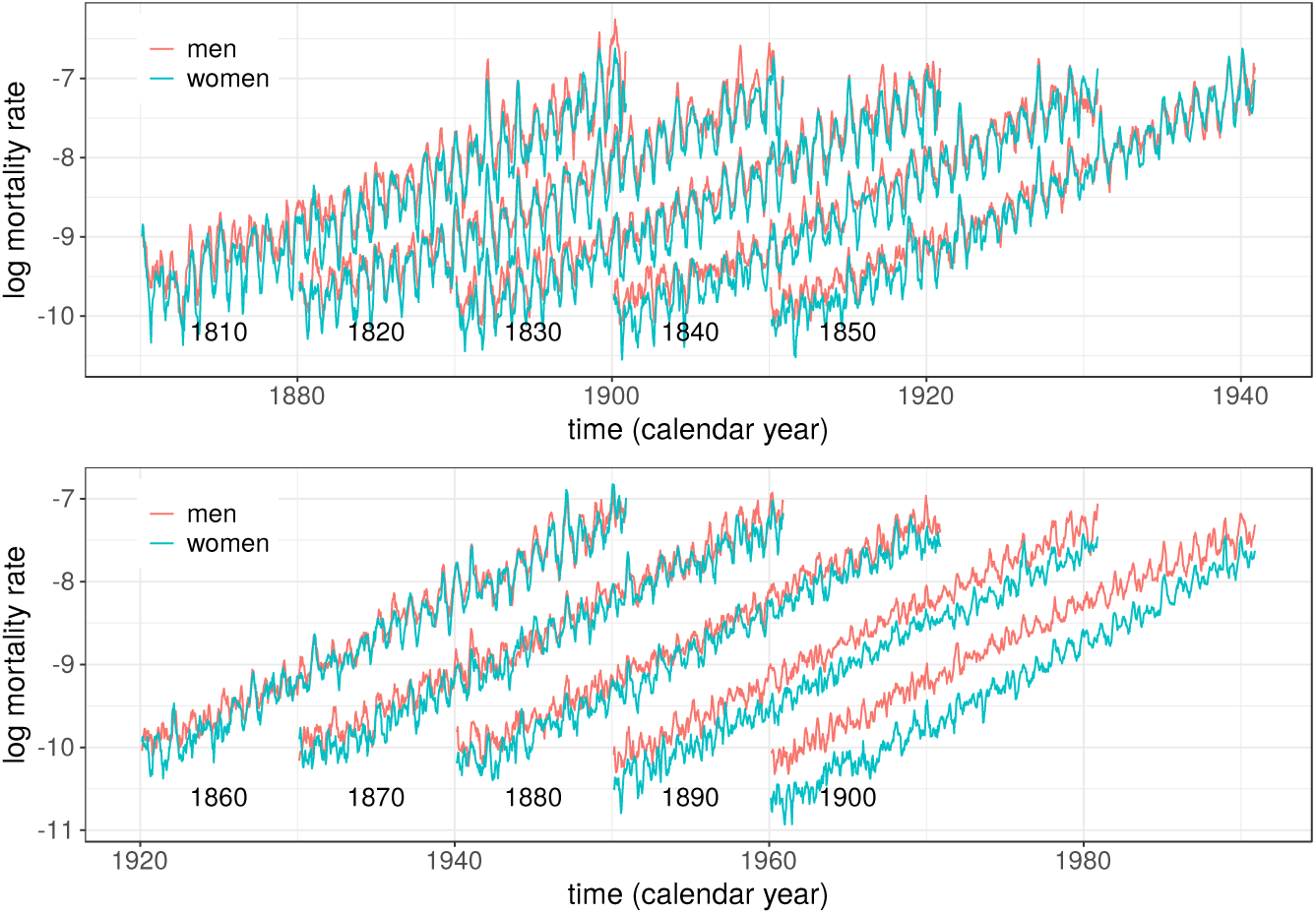
Mortality rates as a function of calendar time for ten birth cohorts of men and women who were born and died in Sweden. Numbers in plots show cohort birth years. Mortality rates shown are local averages over two month (see methods). Data shown for ages between 60 and 90 years.

**Figure 2:**
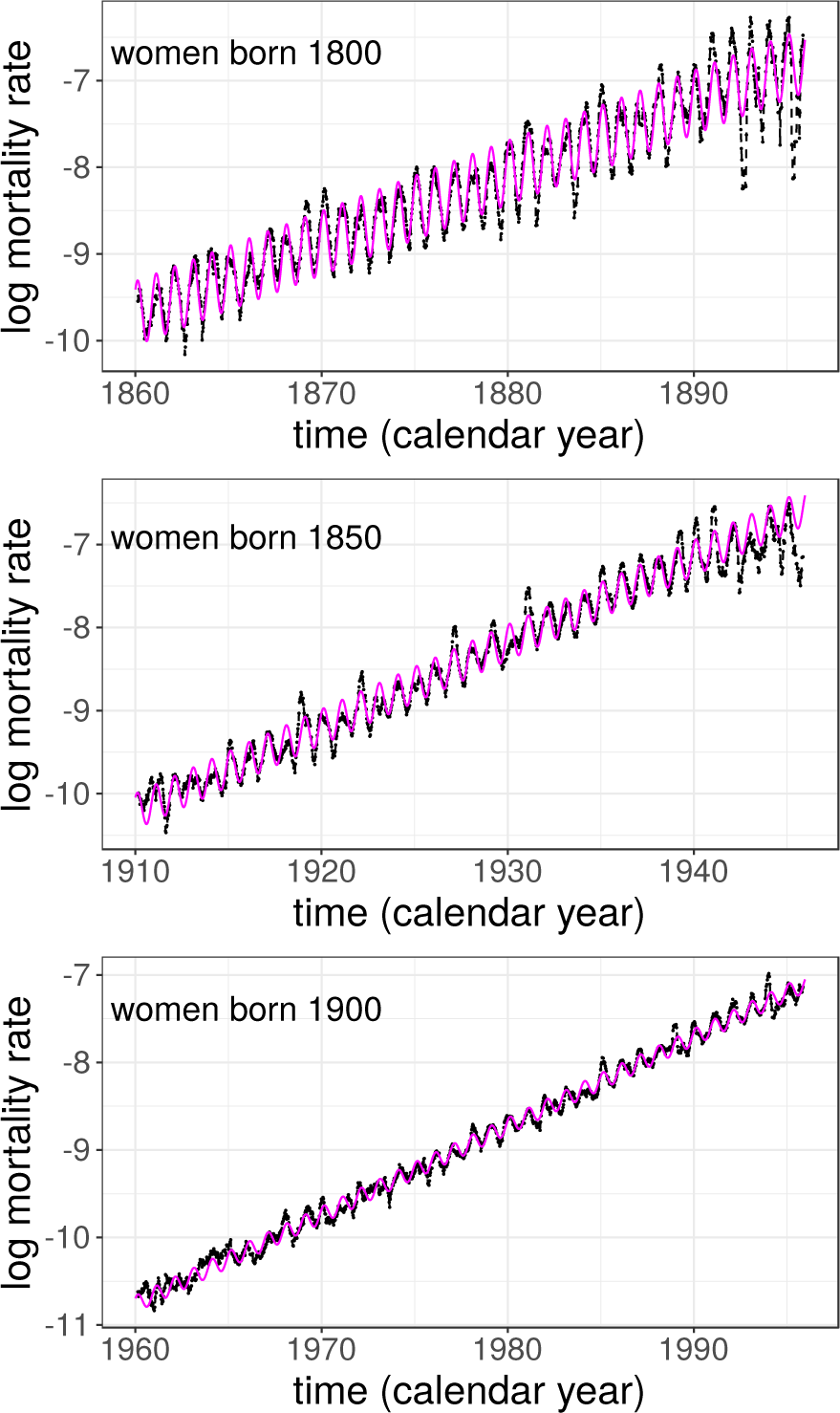
Mortality rates of three birth cohorts of women and predictions from the model (Eq.1) in magenta. Dates on the x-axis (calendar years) indicate the start of the given year. Data shown for ages 60 to 95 years.

### 2.8 Computations

All computations were made in R (R Core Team, 2016). Data and code needed to reproduce the figures is available upon request.

## 3 Results

### 3.1 Overview

Figure 1 shows mortality rates for 20 of the cohorts. Several points are noticeable: The periodicity of mortality rates is highly synchronized between men and women, and its magnitude decreases substantially between cohorts born 1810 and 1900; women have lower mortality rates than men, in particular during summer and early fall (the troughs); The mortality for both men and women decrease between cohorts born in the beginning of the period and those born in the end, and the decrease is more pronounced in the cohorts of women.

The annual peaks of mortality rates occur during winter (supporting Fig. S2) and for cohorts born before 1875 the peak occurred during the second week of February. The mortality peaks for cohorts born 1875-1901 occur somewhat later, extending into March in some cases (supporting Fig S2). Note that this implies, according to the model, that the troughs in mortality occurred during late summer, in the middle to end of August.

### 3.2 A model of seasonality

Mortality rates were modeled by an exponentially increasing function of age modulated by a sinusoidal term with constant amplitude and phase (Eq. 1). Separate fits were made for each of the 202 cohorts (101 sex-specific birth cohorts). The coefficients of the sinusoidal terms were highly significant in all cohorts (F-test, all p-values < 0.001), indicating that the model with seasonality better accounted for the data than did a model with just an exponential increase with age.

Figure 2 shows examples of mortality rates for three cohorts of women with model fits superimposed. Close inspection of the figure shows that sinusoids with constant amplitudes and phases (i.e., Eq. 1) provide good general description of the seasonality present in these data. However, some years have higher rates during winter than what is predicted from the model (e.g., 1931 in the middle panel, due to an influenza outbreak) or lower rates in the summer (e.g., 1879 and 1881 in the top panel). The phases of the seasonal fluctuations are almost constant from year to year, but in some cases the mortality increase comes earlier than predicted. For example, in 1919, in the middle panel, the peak in mortality is earlier than in the model; the Spanish flu hit Sweden in early October 1918 causing the peak to shift. Also, for ages above 90 years, mortality rates become more variable, and are in some cases less well fit by the model (e.g., the 1800 cohort).

Even if the model does not account for all aspects of the data it does provide a compact description of the main trends and provides a low-dimensional summary that will be used below to track changes over time.

### 3.3 Changes in seasonality over time

To illustrate how the magnitude of seasonal variability in mortality has changed over time, the mortality rate ratio (the ratio of the highest to lowest mortality rate in any particular year, Eq. 3) is plotted as a function of cohort birth year in Figure 3. According to the model (Eq. 1) this ratio is constant for a given birth cohort. The magnitude of seasonal variability has decreased substantially over the cohorts, both for men and women. For women born in the first decade of the 19th century, the mortality rate was about two times higher in the winter than in the summer. For women born 90 years later, the ratio had decreased to about 1.1. Also noticeable is that for cohorts born before 1880, the magnitude of seasonality was consistently higher among women than among men.

**Figure 3:**
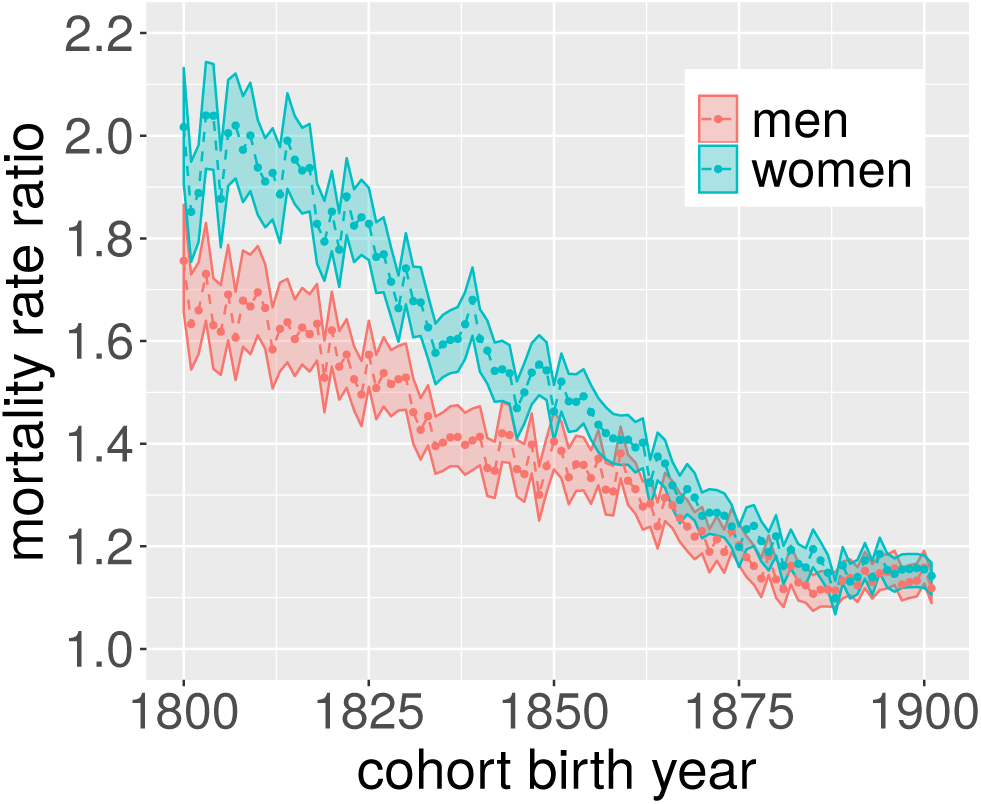
Mortality rate ratios for each cohort were estimated according to Eq. 3 and shows the ratio of the peak of the mortality rates to the trough of the mortality rate for any given year. Colored bands show 95% confidence intervals.

### 3.4 Contribution of seasonality to mean duration of life

To estimate how the decrease in seasonality might have influenced survival, mortality rates were decomposed according to Eq. 4. Figure 4**A** illustrates this decomposition for the cohort of women born 1800. The lower-bound mortality (*M*_1_ in Eq. 4) is seen to provide a good approximation to what the mortality rates would have been if the favorable conditions of late summer would have prevailed throughout the year. Figure 4**B,C** and Tables S2 and S3 show that the cohort life expectancy (conditional upon reaching 59 years of age) increased substantially over the period: about 4.3 years for men and 6.8 years for women. Note that the model fits almost perfectly predicts the life expectancy for these cohorts. For the cohorts of men, Figure 4**B** and Table S2 show that the predicted life expectancy corresponding to the lower-bound mortality increased from 76 years for the cohort born in 1800 to about 78 years for those born 1825, and remained relatively constant in later-born cohorts, indicating that mortality rates during late summer did not decrease further for these cohorts. Figure S3 provides an example of this phenomenon and shows that mortality rates for the 1821 and 1881 cohorts often coincided in the late summers, but differed markedly in winters. Figure 4**C** shows that the survival corresponding to the lower-bound mortality for the cohorts of women increased only slightly over the first 80 cohorts, but over cohorts born between 1880 and 1901 it increased with almost three years.

**Figure 4:**
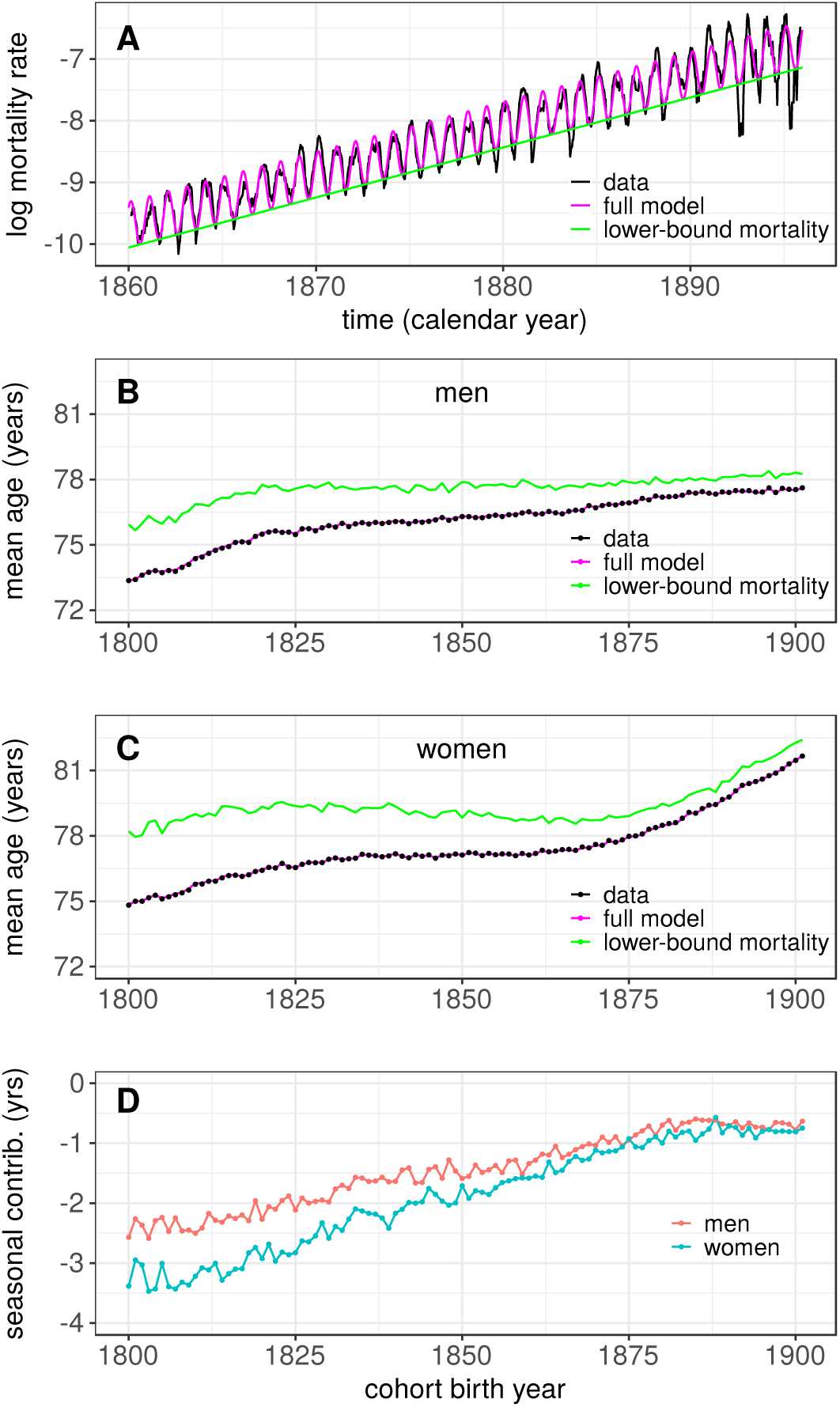
**A**: Illustration of the mortality rate decomposition. Dashed line show mortality data for women born 1800, model fits are shown in magenta. The green line shows the first part, *M*_1_, in the decomposition given by Eq. 4, i.e., the lower-bound mortality. **B**: Life expectancy at 60 for the different cohorts of men. Points show the actual data. The magenta line shows the prediction from the model (Eq. 6). The green line shows the predicted life expectancy, i.e., what life expectancy would have been had mortality rates been equal to the lower-bound mortality (Eq. 4). **C**: same as in **B** but data for the cohorts of women. **D**: Difference between observed mean life duration and mean life duration based on lower-bound mortality in years.

The difference between the actual life expectancy and that predicted based on the lower-bound mortality provides an estimate of the contribution of seasonality and is shown in Figure 4**D**. The effect of seasonality was substantial for cohorts born in the beginning of the 19th century; for women the seasonal variability reduced life expectancy with about 3.5 years, in men by about 2.5. The effect of seasonality first diminished gradually over cohorts, but remained relatively constant at around *−*0.75 years for the last 20 cohorts. This patterns reflects the corresponding decrease in the amplitude of seasonality seen in Fig. 3. Of the total increase in mean life duration, 4.3 years in men and 6.8 in women, this decomposition shows that about 40% of the increase is attributable to the decrease of the magnitude of seasonal fluctuations.

## 4 Discussion

The magnitude of seasonal variations in all-cause mortality rates among elderly in Sweden was shown to have decreased substantially in the cohorts followed (Fig. 3). This is in line with previous results from other countries (Näyhä, 1980; Lerchl, 1998; Kunst et al., 1991; Seretakis et al., 1997; Rau, 2007). Most of these previous studies have focused on secular trends during the second half of the 20th century. The data used here made it possible to show that the decrease in seasonality in Sweden started already with cohorts born during the first decades of the 19th century.

By focusing on cohort data it was possible to quantify the extent to which this reduction of seasonal excess mortality contributed to the increase in cohort life expectancy observed during the time period. Cohort life expectancy at 60 increased by 4.3 years for men and 6.8 years for women, and about 40 percent of this increase could be accounted for by the decrease in seasonality of death.

Below some possible causes of this decrease in seasonality are considered and some technical aspects of the study are discussed.

It should be noted that mortality among younger people, including infants, might also display seasonal variations, sometimes with a different phase compared to older adults, e.g., a mortality peak in the summer (e.g., Rosenwaike, 1966; Näyhä, 1980; Knodel, 1983). It is likely that the mechanisms that drive the seasonal fluctuations in younger people are different from those responsible for the fluctuations among older adults (c.f., Rau, 2007), and the extent to which changes in the magnitude of seasonal variability in mortality among infants and younger people have contributed to changes in life expectancy deserves a separate investigation.

### 4.1 Interpretation of seasonal variation in mortality

Seasonal variability in mortality is a population phenomenon and it is of interest to relate this phenomenon to processes at the individual level. In a recent model of aging and death the exponential increase in mortality with age was shown be a consequence of an accumulation of damage in the individuals (Ledberg, 2020). The rate of accumulation is controlled (in this model) by a damage rate, modeling the amount of damage impinging on the individual, and the rate of repair, i.e., the ability to fix the occurred damage. According to this model, seasonal variability in mortality rates of the type seen here would result if the rate of damage also varied with the seasons. For example, there are several mechanisms through which a decrease in the ambient temperature can increase physiological stress (e.g., Keatinge, 2002), and this might be conceptualized as an increase in the rate of damage, supporting this interpretation. A more indepth investigation of the ability of this model of damage accumulation model and death (Ledberg, 2020) to account for the results reported here is left for a future contribution.

### 4.2 What might have caused the decrease in seasonality

In terms of calendar time, the main part of the decrease in seasonal variability happened between 1870 and 1970, a time-period during which Sweden, as many other countries, went through dramatic social and economical transformations (e.g., Schön, 2010). In 1870 Sweden was a relatively poor country where most people lived on the country side and where about 70% were working in agriculture and forestry. There was little or no public support for old and poor people. In 1970 Sweden had become one the richest countries in the world, the majority of the population lived in cities, less than 10% worked in agriculture, and Sweden had a highly developed welfare system (e.g., Lundberg and Åmark, 2010; Schön, 2010). It is likely that at least part of the decrease in seasonality was caused by the dramatic improvements in living conditions that were part of this transformation of social and economical circumstances (c.f., Kunst et al., 1991). The relative effect of seasonality was found to be approximately constant within birth cohorts and this is an indication of that improvements in living conditions, measured cross-sectionally, are more reflecting conditions among younger generations than among older ones. However, more research is needed to quantify the extent to which improved living conditions played a role in quenching the seasonal variability in mortality.

Another possible explanation of the decrease in seasonality is that the major causes of death might have changed during the time period (c.f. Omran, 1971). Clearly, a change from causes of death more sensitive to the seasons (e.g., respiratory infections) to those less sensitive (e.g., cancers) could lead to a reduction in seasonality similarly to the one observed. However, it is not clear to what extent there has been such a transition in causes of death among older people in Sweden. The system used to classify causes of death has changed multiple times over the time period, and according to official records, less than 10 percent of deaths among people 60 years or older were attributable to infectious diseases already in 1911, a year with substantial seasonal variability. More research is needed to determine to what extent the causes of death among older people in Sweden have changed over this time period.

### 4.3 Differences between men and women

Figure 3 shows that for cohorts born during the first half of the 19th century the seasonal variation in mortality was much more pronounced among women than that among men. Close inspection of Fig. 1 shows that mortality rates among cohorts of women were in general lower than among cohorts of men, and that the difference in seasonality between sexes to a large extent is due to the deeper troughs during the summer and early fall in cohorts of women. One possible explanation of this increased difference in mortality rates during the warm months is that men and women died from different causes. For example, men were much more likely to die from drowning and suicide, two causes of death that show a reverse seasonality and has a peak during the warm months. A quantitative estimate of the extent to which differences in causes of deaths can explain the differences in seasonality would require detailed information about causes of death, something that unfortunately is not available at the national level in Sweden before 1911.

Another striking difference between men and women concerns the development of life expectancy at 60 is shown in Fig. 4. Among women, life expectancy at 60 increased by 3.7 years between cohorts born 1875 and 1901 (Tab. S3). The corresponding increase among men was 0.7 years. Note that this difference is independent of seasonality which was of similar magnitude for men and women during this time period (Fig. 3). The reason for why life expectancy did not increase in men to the same extent might be attributed to sex differences in tobacco smoking, as pointed out previously (Ledberg, 2020). Cigarette smoking became widespread in Sweden during the 1940s and was to start with a predominately male behavior (Svenska Gallup Institutet AB, 1955; Socialstyrelsen, 1986). According to this explanation, life expectancy of men increased less because primarily men were exposed to the deleterious health consequences of smoking. Note that this explanation is consistent with there being no sex differences in the magnitude of seasonality over the concerned time period; indeed the main causes of death related to smoking are relatively independent of the seasons. More research is needed to quantify the contribution of smoking and to identify additional sources that can explain why women saw such an improvement in life expectancy while men did not.

### 4.4 Cohort vs period effects

In this work, seasonal variations in mortality were assessed in a number of extinct birth cohorts. Most previous work on seasonality has been based on period data, and that might seem a more natural choice of data organization. However, given that the magnitude of seasonality was found to be relatively constant within cohorts, the cohort-based analysis provided a very succinct description of the temporal trends (Fig. 3). A major advantage of using cohort data is the possibility to directly compare the magnitude of seasonality to the average life duration.

The model of mortality (Eq. 1) assumes a constant amplitude and phase of seasonal variability in a given birth cohort. This model captured the main trends in the data but it is clear that both the amplitude and phase of seasonality may also depend on calendar year. For example, in years with more pronounced outbreaks of influenza, both amplitudes and phases were affected in multiple cohorts. A more comprehensive account of seasonality should include effects of calendar years as well. However, introducing fixed effects of calendar years in the model only had minor effects on the estimated decrease in seasonality (not shown). For some cohorts, a model where the amplitude of the seasonal effects was allowed to increase with time (i.e., an interaction) provided a better fit to the data. However, the interaction effects were never very strong, and it was judged that the model without interaction captured the magnitude of seasonality well enough for the comparison across cohorts.

The magnitude of seasonal variations has been shown to increase with age in some previous studies (e.g., Näyhä, 1980; Feinstein, 2002; Parks et al., 2018), in the present study, however, the effects were shown to be relatively constant with age (e.g., Fig. 2), and instead to change between birth cohorts. Given that previous works were based on period data, these findings are in fact not contradictory. Indeed, the cohort-dependent effects reported here would show up as age-dependent effects in a period based analysis (as younger age groups belong to later-born cohorts).

### 4.5 Effect of decreasing seasonality

To estimate how much the decrease in seasonality contributed to the increase in average life duration, a comparison was made between the real cohort life expectancy, and the predicted life expectancy based on the lower-bound mortality. This comparison showed that about 40% of the increase in life expectancy at 60 could be attributed to the decrease in seasonality. The interpretability of this estimate depends on that the decomposition of total mortality (Eq. 4) is sensible. Close inspection of Fig 4 shows that the full model almost perfectly predicted cohort life expectancy. Consequently, in terms of life expectancy, the decomposition divided the observed data into two parts with very little left unaccounted for. The data also gave support for that the two parts of the decomposition could change independently of each other, further supporting the sensibility of the decomposition. For example, in supporting Fig. S3 the mortality rates of men born in 1821 are compared to rates of men born in 1881. This figure shows that the differences in mortality rates between these two birth cohort are primarily due to higher rates during winter seasons in the cohort born 1821. The rates during summer seasons are similar in the two cohorts. In terms of life expectancy at 60, those born in 1821 on average lived to be 75.6 whereas those born 1881 lived until 77.2 years. This difference is fully explained by the larger seasonal variability in the first cohort. This example thus shows an increase in life expectancy that was caused entirely by the quenching of the excess mortality in winter seasons, providing additional support for the approach taken here.

Future work might explore in more detail the conditions that have enabled an increased life expectancy among old people in Sweden. The results presented here suggests that a substantial part of this increase might be due to that living conditions in Sweden have become independent of the seasons. However, it is also clear that the bulk part of the increase in life expectancy at 60 must be attributed to factors that are relatively independent of the seasons.

## 5 Conclusions

By following a large number of extinct birth cohorts it was possible to show how the decrease in seasonal fluctuations in mortality has contributed to the increase in average life duration. The simple model of cohort mortality and the decomposition of total mortality introduced here might be useful to better understand the processes and mechanisms underlying the marked improvements in life expectancy seen over the last 150 years.

## Data Availability

Data and code is available upon request.

## 6 Acknowledgements

I thank Sveriges Släktforskarförbund for letting me use data from Dödboken, and an anonymous reviewer for helpful comments.

## Footnotes

1 Explicit reference to a particular cohort (index *j* above) is suppressed for clarity, but all expressions are assumed cohort specific.

