## Supporting information for "A large decrease in the magnitude of seasonal fluctuations in mortality explains part of the increase in longevity in Sweden during 20th century"

Anders Ledberg*

Department of Public Health Sciences, Stockholm University
SE-106 91 Stockholm, Sweden

October 10, 2020

*

### 1 Approximation by Poisson regression

To derive Eq. 2 in the main text a number of approximations were made and here the effect of these approximations are discussed. To measure time in days instead of continuously has no consequences for the age range used in this study; mortality rates change on a much slower time scale.

The expected number of deaths on day $t$ was approximated as

$$E(d_t) = N_t\lambda(t).$$

Note that the correct expression is $E(d_t) = (1 - e^{-\lambda})N$, and the error in approximating $(1-e^{-\lambda})$ by $\lambda$ is quadratic in $\lambda$ by Taylor's theorem. Consequently, this is an excellent approximation when $\lambda(t)$ is small. Recall that $\lambda(t)$ is the mortality rate on day $t$ and note that even at very advanced ages, were the one-year survival probability is some small number, 0.1 say, $\lambda(t)$ will still not be bigger than 0.01. That is, $d_t$ will be well approximated by a Poisson random variable with expected value $N_t\lambda(t)$ throughout.

The final approximation concerns the day of birth for person $i$, that is $b_i$ in Eq. 1 in the main text. To derive Eq. 2 it was tacitly assumed that all persons belonging to the same cohort were born on the same day, in exactly the middle of the year. This assumption made it possible to use the same mortality rate for all people belonging to the same cohort. Of course, people are born throughout the year, and using Eq. 2 to estimate parameters of Eq. 1 might consequently introduce a small bias in the estimates. Note that for $t$ fixed, Eq. 2 represent the average log mortality rate for those still alive at $t$, and those born early in the year will have a slight tendency of dying before those born later in the year (due to the age difference), possibly introducing a bias. The magnitude of this bias was investigated by comparing estimates obtained through fitting Eq. 2 to the data using Poisson regression with estimates obtained by a direct maximization of the likelihood corresponding to Eq. 1 and were found to be negligible (see supporting figure S1). Parameter estimation based on Eq. 2 is preferred because it can be done swiftly using existing routines, and it can moreover be easily extended to incorporate other time-varying factors.

### 2 Figures

#### 2.1 Parameter estimates

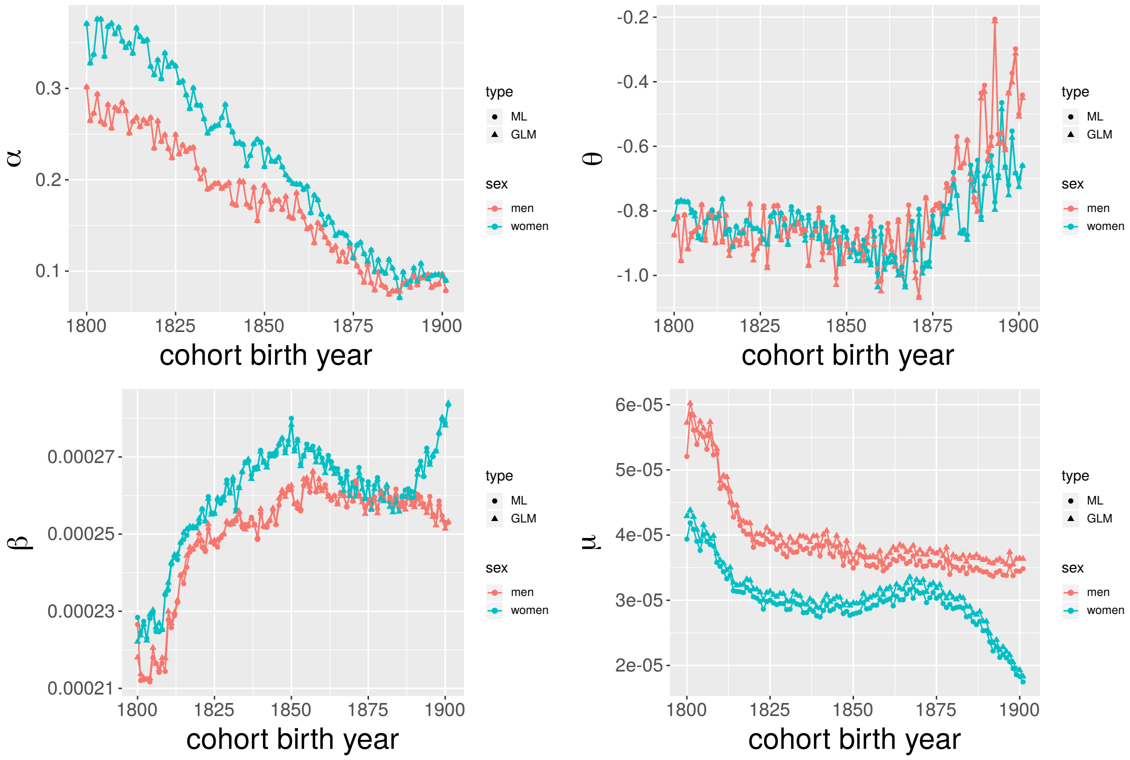

Figure S1: Parameter fits using Poisson regression ("GLM" triangles) and maximum likelihood based on Eq. 1 in the main paper ("ML" circles). The systematic difference in estimates of $\mu$ is due to that the estimates based on Eq. 1 are made for someone 59 years of age at time zero, and the Poisson estimates for someone who is 59.5 years.

#### 2.2 Seasonal location of mortality peak

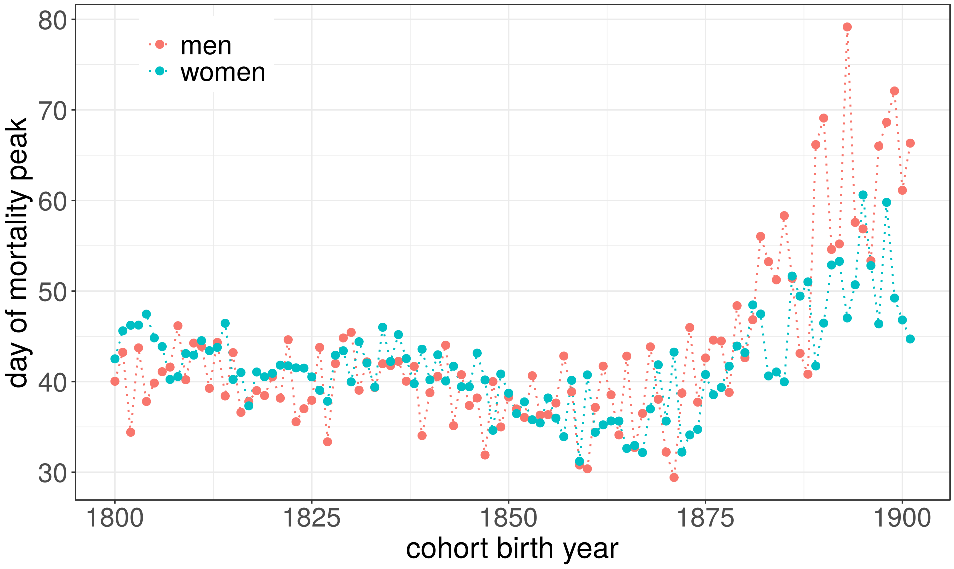

Figure S2: Location of the peak of the seasonal mortality rate fluctuation in days from beginning of the year. The location of the peak was obtained from the model fits.

#### 2.3 Change in the amplitude of seasonality

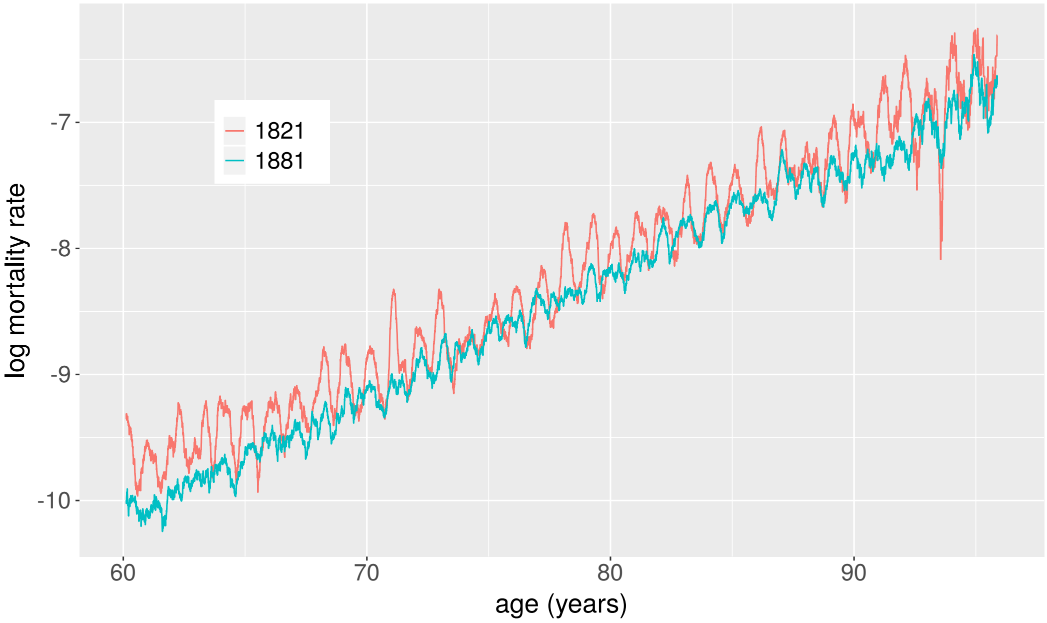

Figure S3: Mortality rates for two birth cohorts of men. Note how the annual troughs often coincide (and are sometimes even lower for the first cohort) but that peaks are consistently higher in the first cohort. This supports the decomposition of total mortality into lower-bound mortality and seasonal mortality. Life expectancy at 60 in the first cohort was 75.6 years compared to 77.2 in the last.

#### 3 Tables

| Birth year | # Men | # Women | Birth year | # Men | # Women |
|---|---|---|---|---|---|
| 1800 | 10316 | 8933 | 1851 | 22489 | 19724 |
| 1801 | 11420 | 9551 | 1852 | 21407 | 18444 |
| 1802 | 12773 | 10769 | 1853 | 22744 | 19803 |
| 1803 | 12351 | 10411 | 1854 | 24903 | 21572 |
| 1804 | 12545 | 10651 | 1855 | 23331 | 20394 |
| 1805 | 12530 | 10652 | 1856 | 23452 | 20063 |
| 1806 | 12014 | 10442 | 1857 | 24362 | 20717 |
| 1807 | 12538 | 10729 | 1858 | 26313 | 22680 |
| 1808 | 11762 | 10077 | 1859 | 26741 | 23029 |
| 1809 | 11302 | 9254 | 1860 | 26585 | 22803 |
| 1810 | 13953 | 11870 | 1861 | 24930 | 21279 |
| 1811 | 15234 | 12854 | 1862 | 26251 | 22771 |
| 1812 | 14822 | 12480 | 1863 | 26806 | 23554 |
| 1813 | 13020 | 11005 | 1864 | 27289 | 23789 |
| 1814 | 14835 | 12589 | 1865 | 26681 | 23657 |
| 1815 | 16633 | 14226 | 1866 | 27056 | 24182 |
| 1816 | 17243 | 14674 | 1867 | 24706 | 21715 |
| 1817 | 16563 | 14218 | 1868 | 22269 | 19232 |
| 1818 | 17232 | 14940 | 1869 | 24069 | 21782 |
| 1819 | 16964 | 14831 | 1870 | 24803 | 22685 |
| 1820 | 17721 | 15405 | 1871 | 26511 | 24464 |
| 1821 | 19237 | 17040 | 1872 | 25821 | 24154 |
| 1822 | 20337 | 17375 | 1873 | 26920 | 24994 |
| 1823 | 21063 | 18616 | 1874 | 27345 | 25571 |
| 1824 | 20322 | 17450 | 1875 | 28234 | 27109 |
| 1825 | 21542 | 18654 | 1876 | 28501 | 27334 |
| 1826 | 20852 | 17887 | 1877 | 30084 | 28713 |
| 1827 | 18571 | 16238 | 1878 | 29555 | 27754 |
| 1828 | 20108 | 17220 | 1879 | 31155 | 29081 |
| 1829 | 20987 | 18256 | 1880 | 30322 | 28286 |
| 1830 | 20130 | 17342 | 1881 | 30411 | 28646 |
| 1831 | 18701 | 16318 | 1882 | 31120 | 28986 |
| 1832 | 19902 | 17466 | 1883 | 31825 | 29143 |
| 1833 | 22048 | 19303 | 1884 | 33288 | 31270 |
| 1834 | 22198 | 19253 | 1885 | 33584 | 31473 |
| 1835 | 21889 | 18925 | 1886 | 34618 | 32250 |
| 1836 | 21267 | 18061 | 1887 | 35420 | 32844 |
| 1837 | 20270 | 17602 | 1888 | 35422 | 32744 |
| 1838 | 19726 | 16761 | 1889 | 34697 | 31691 |
| 1839 | 20291 | 17042 | 1890 | 35793 | 32683 |
| 1840 | 21732 | 18788 | 1891 | 36838 | 33888 |
| 1841 | 20906 | 17883 | 1892 | 36219 | 33691 |
| 1842 | 22125 | 18723 | 1893 | 37396 | 34988 |
| 1843 | 21593 | 18591 | 1894 | 37994 | 35312 |
| 1844 | 22721 | 19759 | 1895 | 39235 | 37056 |
| 1845 | 21619 | 18499 | 1896 | 39465 | 37567 |
| 1846 | 20177 | 17533 | 1897 | 39756 | 38173 |
| 1847 | 20862 | 18025 | 1898 | 40954 | 38987 |
| 1848 | 22087 | 18700 | 1899 | 40727 | 38719 |
| 1849 | 23589 | 20301 | 1900 | 43108 | 40948 |
| 1850 | 22720 | 19583 | 1901 | 43583 | 42030 |

Table S1: Size of birth cohorts

| Birth year | $e_{59}$ | $e_{59}^*$ | diff | Birth year | $e_{59}$ | $e_{59}^*$ | diff |
|---|---|---|---|---|---|---|---|
| 1800 | 73.4 | 75.9 | -2.6 | 1851 | 76.3 | 77.8 | -1.6 |
| 1801 | 73.4 | 75.7 | -2.3 | 1852 | 76.3 | 77.6 | -1.4 |
| 1802 | 73.6 | 76.0 | -2.4 | 1853 | 76.2 | 77.7 | -1.5 |
| 1803 | 73.7 | 76.3 | -2.6 | 1854 | 76.3 | 77.8 | -1.4 |
| 1804 | 73.8 | 76.1 | -2.3 | 1855 | 76.4 | 77.7 | -1.4 |
| 1805 | 73.7 | 76.0 | -2.2 | 1856 | 76.3 | 77.8 | -1.5 |
| 1806 | 73.8 | 76.3 | -2.5 | 1857 | 76.4 | 77.7 | -1.3 |
| 1807 | 73.8 | 76.0 | -2.2 | 1858 | 76.4 | 77.7 | -1.3 |
| 1808 | 74.0 | 76.4 | -2.5 | 1859 | 76.5 | 78.0 | -1.5 |
| 1809 | 74.1 | 76.5 | -2.4 | 1860 | 76.5 | 77.9 | -1.3 |
| 1810 | 74.4 | 76.9 | -2.5 | 1861 | 76.4 | 77.7 | -1.3 |
| 1811 | 74.4 | 76.9 | -2.4 | 1862 | 76.4 | 77.6 | -1.2 |
| 1812 | 74.6 | 76.8 | -2.2 | 1863 | 76.5 | 77.7 | -1.2 |
| 1813 | 74.8 | 77.0 | -2.3 | 1864 | 76.5 | 77.5 | -1.1 |
| 1814 | 74.8 | 77.2 | -2.3 | 1865 | 76.4 | 77.7 | -1.2 |
| 1815 | 74.9 | 77.1 | -2.2 | 1866 | 76.5 | 77.7 | -1.2 |
| 1816 | 75.1 | 77.4 | -2.3 | 1867 | 76.6 | 77.7 | -1.1 |
| 1817 | 75.1 | 77.3 | -2.2 | 1868 | 76.6 | 77.6 | -1.1 |
| 1818 | 75.1 | 77.4 | -2.3 | 1869 | 76.8 | 77.8 | -1.0 |
| 1819 | 75.4 | 77.4 | -2.0 | 1870 | 76.7 | 77.7 | -1.0 |
| 1820 | 75.5 | 77.8 | -2.3 | 1871 | 76.8 | 77.7 | -0.9 |
| 1821 | 75.6 | 77.6 | -2.1 | 1872 | 76.9 | 77.8 | -1.0 |
| 1822 | 75.6 | 77.7 | -2.1 | 1873 | 76.8 | 77.7 | -0.9 |
| 1823 | 75.6 | 77.5 | -2.0 | 1874 | 76.9 | 77.9 | -1.0 |
| 1824 | 75.6 | 77.5 | -1.9 | 1875 | 76.9 | 77.9 | -0.9 |
| 1825 | 75.5 | 77.6 | -2.1 | 1876 | 77.0 | 77.8 | -0.9 |
| 1826 | 75.7 | 77.7 | -1.9 | 1877 | 77.1 | 77.9 | -0.8 |
| 1827 | 75.7 | 77.7 | -2.0 | 1878 | 77.1 | 77.8 | -0.7 |
| 1828 | 75.6 | 77.6 | -2.0 | 1879 | 77.2 | 78.1 | -0.9 |
| 1829 | 75.8 | 77.7 | -1.9 | 1880 | 77.2 | 77.9 | -0.7 |
| 1830 | 75.9 | 77.9 | -2.0 | 1881 | 77.2 | 77.8 | -0.6 |
| 1831 | 75.8 | 77.6 | -1.8 | 1882 | 77.2 | 78.0 | -0.8 |
| 1832 | 76.0 | 77.7 | -1.7 | 1883 | 77.3 | 77.9 | -0.7 |
| 1833 | 75.8 | 77.6 | -1.8 | 1884 | 77.4 | 78.1 | -0.6 |
| 1834 | 76.0 | 77.5 | -1.6 | 1885 | 77.4 | 78.0 | -0.6 |
| 1835 | 76.0 | 77.6 | -1.6 | 1886 | 77.4 | 78.1 | -0.6 |
| 1836 | 76.0 | 77.6 | -1.6 | 1887 | 77.4 | 78.0 | -0.6 |
| 1837 | 76.0 | 77.7 | -1.6 | 1888 | 77.3 | 77.9 | -0.6 |
| 1838 | 76.0 | 77.5 | -1.6 | 1889 | 77.4 | 78.1 | -0.7 |
| 1839 | 76.0 | 77.7 | -1.6 | 1890 | 77.4 | 78.1 | -0.7 |
| 1840 | 76.1 | 77.7 | -1.6 | 1891 | 77.5 | 78.1 | -0.6 |
| 1841 | 76.1 | 77.5 | -1.4 | 1892 | 77.5 | 78.2 | -0.7 |
| 1842 | 76.0 | 77.4 | -1.4 | 1893 | 77.5 | 78.2 | -0.7 |
| 1843 | 76.1 | 77.7 | -1.7 | 1894 | 77.4 | 78.2 | -0.7 |
| 1844 | 76.0 | 77.7 | -1.7 | 1895 | 77.4 | 78.2 | -0.7 |
| 1845 | 76.1 | 77.5 | -1.4 | 1896 | 77.6 | 78.4 | -0.8 |
| 1846 | 76.1 | 77.5 | -1.4 | 1897 | 77.4 | 78.1 | -0.7 |
| 1847 | 76.3 | 77.8 | -1.6 | 1898 | 77.6 | 78.3 | -0.7 |
| 1848 | 76.1 | 77.4 | -1.3 | 1899 | 77.6 | 78.2 | -0.7 |
| 1849 | 76.2 | 77.7 | -1.5 | 1900 | 77.5 | 78.3 | -0.8 |
| 1850 | 76.3 | 77.9 | -1.6 | 1901 | 77.6 | 78.3 | -0.6 |

Table S2: Survival data for men. $e_{59}$ is the average life duration conditional upon reaching 59.5 years of age. $e_{59}^*$ is the prediction from the model based on the lower-bound mortality, that is $M_1$ in Equation 4 in the main paper.

| Birth year | $e_{59}$ | $e^*_{59}$ | diff | Birth year | $e_{59}$ | $e^*_{59}$ | diff |
|---|---|---|---|---|---|---|---|
| 1800 | 74.8 | 78.2 | -3.4 | 1851 | 77.2 | 79.2 | -1.9 |
| 1801 | 75.0 | 78.0 | -2.9 | 1852 | 77.2 | 79.0 | -1.8 |
| 1802 | 75.0 | 78.0 | -3.0 | 1853 | 77.1 | 78.9 | -1.8 |
| 1803 | 75.2 | 78.6 | -3.5 | 1854 | 77.2 | 79.1 | -1.9 |
| 1804 | 75.3 | 78.7 | -3.4 | 1855 | 77.1 | 78.9 | -1.7 |
| 1805 | 75.1 | 78.1 | -3.0 | 1856 | 77.2 | 78.8 | -1.7 |
| 1806 | 75.2 | 78.6 | -3.4 | 1857 | 77.2 | 78.8 | -1.6 |
| 1807 | 75.3 | 78.7 | -3.4 | 1858 | 77.1 | 78.7 | -1.6 |
| 1808 | 75.4 | 78.7 | -3.3 | 1859 | 77.2 | 78.8 | -1.6 |
| 1809 | 75.5 | 78.9 | -3.4 | 1860 | 77.1 | 78.7 | -1.6 |
| 1810 | 75.8 | 79.0 | -3.2 | 1861 | 77.2 | 78.7 | -1.5 |
| 1811 | 75.8 | 78.9 | -3.1 | 1862 | 77.3 | 78.9 | -1.6 |
| 1812 | 75.9 | 79.0 | -3.1 | 1863 | 77.3 | 78.6 | -1.3 |
| 1813 | 75.9 | 78.9 | -3.0 | 1864 | 77.3 | 78.8 | -1.5 |
| 1814 | 76.1 | 79.4 | -3.3 | 1865 | 77.4 | 78.8 | -1.5 |
| 1815 | 76.2 | 79.4 | -3.2 | 1866 | 77.4 | 78.7 | -1.3 |
| 1816 | 76.2 | 79.3 | -3.1 | 1867 | 77.3 | 78.6 | -1.2 |
| 1817 | 76.1 | 79.2 | -3.1 | 1868 | 77.5 | 78.8 | -1.3 |
| 1818 | 76.2 | 79.0 | -2.8 | 1869 | 77.4 | 78.7 | -1.3 |
| 1819 | 76.4 | 79.1 | -2.7 | 1870 | 77.6 | 78.7 | -1.1 |
| 1820 | 76.4 | 79.3 | -2.9 | 1871 | 77.6 | 78.7 | -1.2 |
| 1821 | 76.6 | 79.2 | -2.7 | 1872 | 77.8 | 78.9 | -1.1 |
| 1822 | 76.5 | 79.5 | -3.0 | 1873 | 77.7 | 78.8 | -1.1 |
| 1823 | 76.7 | 79.6 | -2.8 | 1874 | 77.8 | 78.9 | -1.1 |
| 1824 | 76.6 | 79.4 | -2.9 | 1875 | 78.0 | 78.9 | -0.9 |
| 1825 | 76.5 | 79.4 | -2.8 | 1876 | 78.0 | 79.1 | -1.1 |
| 1826 | 76.7 | 79.3 | -2.6 | 1877 | 78.1 | 79.2 | -1.1 |
| 1827 | 76.8 | 79.4 | -2.6 | 1878 | 78.3 | 79.3 | -1.0 |
| 1828 | 76.8 | 79.3 | -2.5 | 1879 | 78.4 | 79.3 | -0.9 |
| 1829 | 76.8 | 79.1 | -2.3 | 1880 | 78.5 | 79.5 | -1.0 |
| 1830 | 76.9 | 79.5 | -2.6 | 1881 | 78.6 | 79.4 | -0.8 |
| 1831 | 77.0 | 79.4 | -2.4 | 1882 | 78.6 | 79.5 | -0.9 |
| 1832 | 76.9 | 79.4 | -2.5 | 1883 | 78.8 | 79.6 | -0.8 |
| 1833 | 76.9 | 79.2 | -2.3 | 1884 | 79.1 | 79.9 | -0.8 |
| 1834 | 77.0 | 79.1 | -2.1 | 1885 | 79.0 | 80.0 | -1.0 |
| 1835 | 77.2 | 79.3 | -2.1 | 1886 | 79.3 | 80.1 | -0.8 |
| 1836 | 77.1 | 79.3 | -2.2 | 1887 | 79.4 | 80.2 | -0.8 |
| 1837 | 77.1 | 79.3 | -2.2 | 1888 | 79.4 | 80.0 | -0.6 |
| 1838 | 77.0 | 79.3 | -2.2 | 1889 | 79.7 | 80.5 | -0.8 |
| 1839 | 77.1 | 79.5 | -2.4 | 1890 | 79.8 | 80.5 | -0.7 |
| 1840 | 77.2 | 79.3 | -2.2 | 1891 | 80.1 | 80.8 | -0.7 |
| 1841 | 77.1 | 79.2 | -2.1 | 1892 | 80.3 | 81.2 | -0.9 |
| 1842 | 77.0 | 79.0 | -2.0 | 1893 | 80.4 | 81.2 | -0.8 |
| 1843 | 77.1 | 79.1 | -2.0 | 1894 | 80.5 | 81.4 | -0.9 |
| 1844 | 77.1 | 79.0 | -2.0 | 1895 | 80.6 | 81.4 | -0.8 |
| 1845 | 77.1 | 78.9 | -1.8 | 1896 | 80.8 | 81.5 | -0.8 |
| 1846 | 77.0 | 78.8 | -1.9 | 1897 | 80.9 | 81.7 | -0.8 |
| 1847 | 77.1 | 79.1 | -2.0 | 1898 | 81.1 | 81.9 | -0.8 |
| 1848 | 77.1 | 79.1 | -2.0 | 1899 | 81.3 | 82.1 | -0.8 |
| 1849 | 77.2 | 79.2 | -2.0 | 1900 | 81.5 | 82.3 | -0.8 |
| 1850 | 77.1 | 78.8 | -1.7 | 1901 | 81.7 | 82.4 | -0.7 |

Table S3: Survival data for women. $e_{59}$ is the average life duration conditional upon reaching 59.5 years of age. $e^*_{59}$ is the prediction from the model based on the lower-bound mortality, that is $M_1$ in Equation 4 in the main paper.
